# Plant-based fat supply is associated with reduced ADHD disease burden

**DOI:** 10.1101/2024.01.30.24301956

**Authors:** Duan Ni, Alistair Senior, David Raubenheimer, Stephen J. Simpson, Ralph Nanan

**Affiliations:** Sydney Medical School Nepean, The University of Sydney, Sydney, NSW, Australia; Nepean Hospital, Nepean Blue Mountains Local Health District, Sydney, NSW, Australia; Charles Perkins Centre, The University of Sydney, Sydney, NSW, Australia; School of Life and Environmental Sciences, The University of Sydney, Sydney, NSW, Australia; Sydney Precision Data Science Centre, The University of Sydney, Sydney, NSW, Australia

## Abstract

**Objectives:** Attention-Deficit/Hyperactivity Disorder (ADHD) is emerging as a major neurodevelopmental disorder on a global scale, affecting both children and increasingly adult population. Its aetiology is unclear but seems to involve genetic and environmental factors, particularly diets and nutrients. However, most studies so far only focused on specific nutrients or dietary patterns, lacking systematic perspectives of their potential interactions, and also neglecting other confounding factors like socioeconomic status. Thus, we aim to systematically interrogate the association between nutrient supply, reflecting the food exposure and environment, socioeconomic status and ADHD disease burden at a global level over time.

**Methods:** ADHD disease burden, macronutrient supply and gross domestic product (GDP) were collated from more than 150 countries from 1990 to 2018 and analyzed with nutritional geometry generalized additive mixed models (GAMMs).

**Results:** Modelling results suggested the interactive effects of nutrients and socioeconomic status on ADHD. Fat, especially plant-based fat supply, is associated with decreased ADHD disease burden. These associations were conserved across sexes and ages and were not confounded by the total energy supply.

**Conclusions:** Globally, far, particularly plant-based fat supply seemed to drive the reduction of ADHD disease burden, which is supported by previous reports about the amelioration of ADHD by ketogenic diets. Further in-depth studies are needed to elucidate the underlying mechanistic and may inform future targeted dietary interventions for ADHD prevention and/or treatment.

## Introduction

Attention-Deficit/Hyperactivity Disorder (ADHD) is a neurodevelopmental disorder, with a peak prevalence in childhood and a gender bias towards males^1^. Its aetiology is still unclear but appears to involve both genetic and environmental factors. Diets have been a focus of interest and some studies suggested “Western diets” aggravate, while Mediterranean diets ameliorate ADHD^2^. However, previous dietary research yielded mixed results, possibly due to focusing on single nutritional factors while neglecting broader socioeconomic contexts and their plausible interactions.

Here, we harnessed nutritional geometry generalized additive mixed models (GAMMs)^3^, to model the associations between macronutrient supply, reflecting population food exposures, and ADHD burden globally, whilst correcting for socioeconomic status and their potential interactions. Our analyses showed that increased fat, particularly plant-based fat supply, is linked to less ADHD. This will warrant future mechanistic studies to unravel the interplay between macronutrients and ADHD, and potentially pave the way towards targeted nutritional interventions against ADHD.

## Methods

ADHD data is available from Global Burden of Disease website (http://ghdx.healthdata.org) and nutrient supply and GDP data was obtained as previously described^3,4^. Analysis details are described in Supplementary Information and adapted from previous works^3,4^. In brief, GAMMs were used to analyze the effects of nutrient supplies and GDP over time on ADHD data globally.

## Results

Global age-standardized ADHD prevalence and incidence mildly decreased over time (Figure 1A), accompanied by increases in global nutrient supplies and GDP (Figure 1B). These parameters are potentially interconnected, posing challenges to delineating their detailed contributions and associations (Figure 1C-E). Hence, GAMMs were deployed to interrogate their associations with ADHD. A model considering the interactions between nutrient supplies and GDP, adding the time effects, was favoured (Supplementary Information), suggesting the interactive effects of nutrients and socioeconomic status on ADHD.

**Figure 1.**
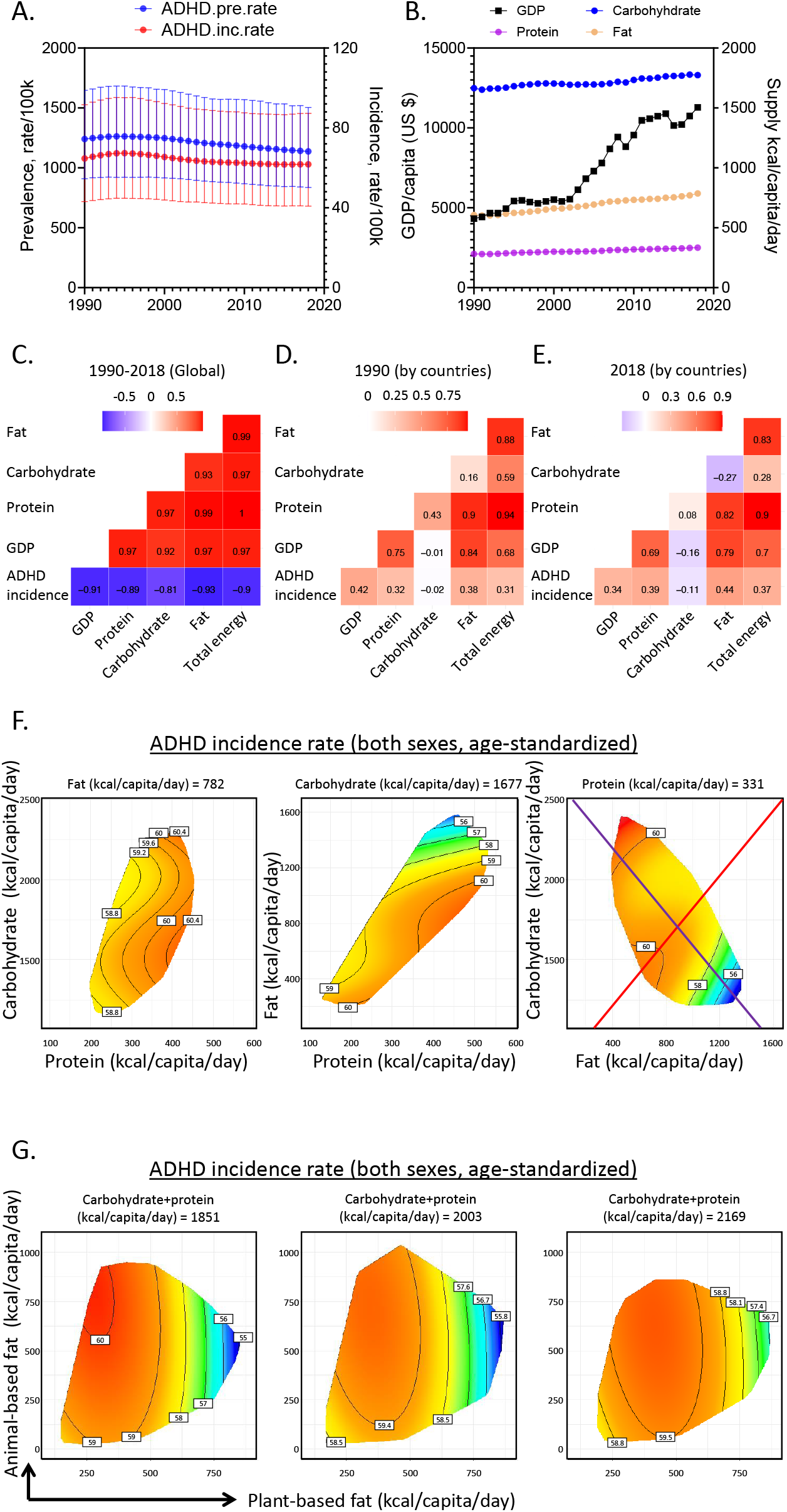
Association of global macronutrient supply and Attention-Deficit/Hyperactivity Disorder (ADHD) disease burden. **A**. Global age-standardized ADHD prevalence (blue) and incidence rate (red) of both sexes as functions of year. **B**. Global GDP per capita (in US dollars, black) and supplies of carbohydrate (blue), protein (purple) and fat (brown) as functions of year. **C-E**. Correlations for global variables from 1990-2018 (**C**) and for variables among different countries in 1990 (**D**) and 2018 (**E**). Correlation coefficients are shown. **F**. Predicted effects of macronutrient supply on age-standardized ADHD incidence rate of both sexes. The left panel shows the effect of carbohydrate and protein supply, the middle shows fat and protein and the right shows carbohydrate and fat supplies. In each panel, the third macronutrient is set at median level (value shown). Surface colors reflect the modelled results, with red indicates higher ADHD incidence and blue means lower. **G**. Predicted effects of carbohydrate and protein, animal- and plant-based fat supplies on ADHD incidence. Modelled results were plotted in 2-dimensional nutritional space with plant-based fat supply as *x*-axis and animal-based fat supply as *y*-axis, with supply of carbohydrate and protein held at 25%, 50% and 75% quantiles of global data. (See Supplementary Information for statistics and interpretation).

Figure 1F shows the results of 2018, the most recent year with relatively comprehensive data coverage. Carbohydrate and protein supplies minimally influenced ADHD. In contrast, increasing fat supply strongly correlated with lower ADHD incidence. The purple vector is an isocaloric line, along which the total energy supply is held constant, and fat is isocalorically substituted for carbohydrate. Increasing fat:carbohydrate ratio lowered ADHD incidence. Similar patterns were found for ADHD prevalence and across genders and ages (Supplementary Figure 1). Along the red radial, increasing total energy supply, whilst holding fat:carbohydrate ratio constant, had little impact on ADHD incidence.

Effects of fat were further investigated, by sub-setting total fat into plant- (*x*-axis) and animal-based (*y*-axis) fat supplies while combining carbohydrate and protein supplies (Figure 1G). We showed the effects of plant and animal fats, assuming low, medium and high carbohydrate and protein supplies. Increasing plant-based fat supply robustly decreased ADHD while animal-based fat conferred limited effects.

## Discussion

This represents the first study comprehensively probing the interplay of nutrient supply, as a proxy of food environment, and ADHD. Our results suggest a potential protective role of fat, particularly plant-based fat. Their increasing supplies are associated with reduced ADHD disease burden.

These results align with studies documenting ADHD amelioration by ketogenic diets^5^. The fact that plant-based but not animal-based fat contributed to ADHD reduction in our analysis, is in accord with prior studies demonstrating the benefits of vegetable-enriched diets for ADHD^2^. Similarly, polyunsaturated fatty acids, a major component of plant-based fats, have been reported as co-adjuvant for ADHD treatment^6^. Nevertheless, further researches are needed to demonstrate the potential causality, as well as to interrogate whether plant-based fats exert preventive effects *in utero*, during early infancy and/or directly modify ADHD.

Collectively, we have established a clear link between global nutrient supply, reflecting the food environment, and ADHD disease burden, which might inform future dietary interventions with plant-based fat either for ADHD prevention or treatment.

## Data Availability

All data produced in the present study are available upon reasonable request to the authors

Duan Ni, PhD

Alistair M. Senior, PhD

David Raubenheimer, PhD

Stephen J. Simpson, PhD

Ralph Nanan, Dr. med. Habil (Germany). FRACP

## Author Contributions

*Concept and design:* Duan Ni, and Ralph Nanan.

*Acquisition, analysis, and interpretation of data:* Duan Ni, Alistair M. Senior, David Raubenheimer, Stephen J. Simpson, and Ralph Nanan.

*Drafting of the manuscript:* Duan Ni, and Ralph Nanan.

*Critical revision of the manuscript for important intellectual content:* All authors.

