## Supplementary Information for "Plant-based fat supply is associated with reduced ADHD disease burden"

**Interpretation of modelling surfaces**

Details for modelling surface interpretation are adapted from previous publications^1-3^. In brief, using RStudio (v4.2.2.), Attention-Deficit/Hyperactivity Disorder (ADHD) disease burden data were analyzed with generalized additive mixed models (GAMMs) prediction^4,5^. For analysis of the supplies of three macronutrients, results were mapped as response surfaces based on nutrient axes for three macronutrients, protein, carbohydrate and fat. For analysis of fat types, modelling results were projected on responses with plant-based fat on the x-axis and animal-based fat on the y-axis, while holding carbohydrate and protein supply at a range of constants (i.e., 25%, 50% (median) and 75% quantiles of the global supply of different countries).

Within the modelling surfaces, red reflects higher values, while blue implies lower ones. Along the black contour lines, the modelling values are unchanged and the numbers on the lines note the magnitude of the parameters. The purple line is an isocaloric line, along which the total energy supply from macronutrients is unchanged but fat is substituted for carbohydrate isocalorically. The red line is a food rail. Carbohydrate:fat ratio is held constant along it, but the total macronutrient energy supply is altered.

Statistics for the modelling analysis are provided in Supplementary Tables. When the modelling analysis is significant, the effect of macronutrient supply on the modelling values (i.e., ADHD disease burden) can be deduced from the modelling surfaces.

**Supplementary Methods**

*Data collection and processing*

Data collation and processing were adapted from previous studies^1,3^. ADHD data was obtained from the Global Burden of Disease Study 2019 (GBD 2019). As described^1^, macronutrient supply data and gross domestic product (GDP) data were collected from the Food and Agriculture Organization Corporate Statistical Database (FAOSTAT, [www.fao.org/faostat/en/#home](http://www.fao.org/faostat/en/#home)) and the Maddison project^6^ respectively.

Analyses were based on data from 1990 to 2018 with relatively comprehensive data coverage. Countries or time points with no data record were filtered and the resulting data spanning from 1990 to 2018 covering more than 150 countries were further analyzed with R.

*Generalized additive mixed models (GAMMs)*

Details of GAMMs were described in^1,3^. In brief, GAMMs^3,4^ were used to model the changes in ADHD burden over time and evaluate the impacts from macronutrient supply and GDP. GAMMs take into account the nonlinear terms as nonparametric smoothed functions, often in a form of spline, and provide a flexible manner to estimate the nonlinear associations. All modelling was carried out with the *mgcv* package and its “gam” function^4,5^. All models considered the country that the data were based on as a random effect. The gamma parameter, implying the smoothing degrees of the modelled effects was defined as log(*n*)/2, where *n* is the number of combinations for countries and years with available data. A Gaussian family with log-link function was used for modelling.

Several different predictor variables and their different combinations as well as a null model where only the random effect from the country is considered are compared. Models with multiple variables consider all combinations of the individual, additive and interactions among parameters like macronutrient supply, year and GDP data. Macronutrient supply was modelled as a three-dimensional spline, and year and GDP data were modelled as one-dimensional cubic-regression splines utilizing the “s()” function in *mgcv* package. The interactions between smooth terms on different scales such as macronutrient supply and year were modelled with “te()” function from *mgcv* package using tensor product smoothers.

Modelling results were compared using Akaike information criterions (AICs) and the model with the lowest AIC was selected^7^.

Codes for the analysis are adapted from GitHub, <https://github.com/Nidane/Asthma-NutrientSupply>.

**Supplementary Tables**

Supplementary Tables 1-4 are GAMMs estimates. For parametric terms the model estimates and associated standard errors (SE) and test statistics are presented. For non-parametric smooth terms, the estimated and reference degrees of freedom (reflected by edf, sumEDF, Ref.df) are shown as well as their test statistics. Smooth terms were fitter with either the standard smooth function “s()” or a tensor product smooth “te()” in the *mgcv* package. Tensor product smoothing was utilized where terms exist on different units (for example, nutrient supply and year). Country was included as a random effect using the smooth function “s()”. The model family and implemented link function is stated.

**Supplementary Table 1.** Relative fit of generalized additive mixed models (GAMMs) testing the predictors for age-standardized ADHD incidence rate in both sexes. Gamma is the degrees of freedom inflation factor. Dev means the deviance explained. AIC = Akaike information criterion. GDP = gross domestic product. Delta is the differences between AICs of models and the minimum AIC. sumEDF reflects the degrees of freedom of the models. Macronutrient supply was modelled as a three-dimensional thin-plate spline. (related to Figure 1F)

| **GAM** | **Gamma** | **Dev** | **AIC** | **Delta** | **Weights** | **sumEDF** | **Formula** |
| --- | --- | --- | --- | --- | --- | --- | --- |
| 1 | 3.855439 | 99.76844 | 17867.64 | 1633.088 | 0 | 157.8517 | 1 + s(Country, bs="re") |
| 2 | 3.855439 | 99.77226 | 17813.08 | 1578.518 | 0 | 167.6359 | s(protein.kcal, carb.kcal, fat.kcal, k=k_nut) + s(Country, bs="re") |
| 3 | 3.855439 | 99.78785 | 17481.19 | 1246.629 | 1.98567010450277e-271 | 160.0696 | s(Year, k=10, bs="cr") + s(Country, bs="re") |
| 4 | 3.855439 | 99.79119 | 17420.92 | 1186.361 | 2.42622252865814e-258 | 165.3298 | s(GDP, k=10, bs="cr") + s(Country, bs="re") |
| 5 | 3.855439 | 99.79188 | 17415.36 | 1180.801 | 3.91213781832251e-257 | 169.955 | s(protein.kcal, carb.kcal, fat.kcal, k=k_nut) + s(Year, k=10, bs="cr") + s(Country, bs="re") |
| 6 | 3.855439 | 99.7939 | 17374.45 | 1139.895 | 2.9853797841001e-248 | 171.3127 | s(protein.kcal, carb.kcal, fat.kcal, k=k_nut) + s(GDP, k=10, bs="cr") + s(Country, bs="re") |
| 7 | 3.855439 | 99.79369 | 17371.53 | 1136.97 | 1.28866084850942e-247 | 167.5382 | s(Year, k=10, bs="cr") + s(GDP, k=10, bs="cr") + s(Country, bs="re") |
| 8 | 3.855439 | 99.82545 | 16670.65 | 436.089 | 2.01589199378044e-95 | 190.2406 | te(protein.kcal, carb.kcal, fat.kcal, Year, bs=c("tp", "cr"), d=c(3,1), k=c(k_nut, 7)) + s(Country, bs="re") |
| 9 | 3.855439 | 99.84142 | 16262.38 | 27.8253 | 9.07427843328068e-07 | 200.3362 | te(protein.kcal, carb.kcal, fat.kcal, GDP, bs=c("tp", "cr"), d=c(3,1), k=c(k_nut, 7)) + s(Country, bs="re") |
| 10 | 3.855439 | 99.80421 | 17161.24 | 926.679 | 5.94592930369434e-202 | 179.2737 | te(Year, GDP, k=10) + s(Country, bs="re") |
| 11 | 3.855439 | 99.83117 | 16539.13 | 304.5756 | 7.28191457923261e-67 | 198.9599 | te(protein.kcal, carb.kcal, fat.kcal, Year, bs=c("tp", "cr"), d=c(3,1), k=c(k_nut, 7)) + s(GDP, k=10, bs="cr") + s(Country, bs="re") |
| **12** | **3.855439** | **99.84247** | **16234.56** | **0** | **0.999999** | **201.3021** | **te(protein.kcal, carb.kcal, fat.kcal, GDP, bs=c("tp", "cr"), d=c(3,1), k=c(k_nut, 7)) + s(Year, k=10, bs="cr") + s(Country, bs="re")** |
| 13 | 3.855439 | 99.8091 | 17068.62 | 834.0669 | 7.66797594645443e-182 | 189.414 | te(Year, GDP, k=10) + s(protein.kcal, carb.kcal, fat.kcal, k=k_nut) + s(Country, bs="re") |

**Supplementary Table 2.** Estimated effects of macronutrient supply by time and GDP per capita on age-standardized ADHD incidence rate. Gaussian-GAMM, log-link function. (related to Figure 1F)

| **Parametric coefficients** | |  |  |  |
| --- | --- | --- | --- | --- |
|  | **Estimate** | **Std. Error** | **t value** | **Pr(>\|t\|)** |
| **(Intercept)** | 4.0736 | 0.0304 | 134 | <2e-16 |
| **Approximate significance of smooth terms** | | | |  |
|  | **edf** | **Ref.df** | **F** | **p-value** |
| **te(protein.kcal,carb.kcal,fat.kcal,GDP)** | 42.36 | 47.14 | 29.62 | <2e-16 |
| **s(Year)** | 1.00 | 1.00 | 25.87 | 6.2e-07 |
| **s(Country)** | 157.94 | 158.00 | 11586.14 | <2e-16 |
| R-sq.(adj) = 0.998 Deviance explained = 99.8% | | | | |
| GCV = 2.9772 Scale est. = 2.1242 n = 4465 | | | | |

**Supplementary Table 3.** Relative fit of generalized additive mixed models (GAMMs) testing the predictors for age-standardized ADHD incidence rate in both sexes. Gamma is the degrees of freedom inflation factor. Dev means the deviance explained. pbf = plant-based fat supply. carb_prot = carbohydrate and protein supply. abf = animal-based fat supply. AIC = Akaike information criterion. GDP = gross domestic product. Delta is the differences between AICs of models and the minimum AIC. sumEDF reflects the degrees of freedom of the models. Macronutrient supply (plant- and animal-based fats and carbohydrate and protein) was modelled as a three dimensional thin-plate spline. (related to Figure 1G)

| **GAM** | **Gamma** | **Dev** | **AIC** | **Delta** | **Weights** | **sumEDF** | **Formula** |
| --- | --- | --- | --- | --- | --- | --- | --- |
| 1 | 3.855439 | 99.76844 | 17867.64 | 2022.136 | 0 | 157.8517 | 1 + s(Country, bs="re") |
| 2 | 3.855439 | 99.77581 | 17742.72 | 1897.214 | 0 | 167.6289 | s(pbf.kcal, carb_prot.kcal, abf.kcal, k=k_nut) + s(Country, bs="re") |
| 3 | 3.855439 | 99.78785 | 17481.19 | 1635.677 | 0 | 160.0696 | s(Year, k=10, bs="cr") + s(Country, bs="re") |
| 4 | 3.855439 | 99.79119 | 17420.92 | 1575.409 | 0 | 165.3298 | s(GDP, k=10, bs="cr") + s(Country, bs="re") |
| 5 | 3.855439 | 99.79299 | 17390.87 | 1545.365 | 0 | 169.6712 | s(pbf.kcal, carb_prot.kcal, abf.kcal, k=k_nut) + s(Year, k=10, bs="cr") + s(Country, bs="re") |
| 6 | 3.855439 | 99.79506 | 17349.34 | 1503.827 | 0 | 171.2812 | s(pbf.kcal, carb_prot.kcal, abf.kcal, k=k_nut) + s(GDP, k=10, bs="cr") + s(Country, bs="re") |
| 7 | 3.855439 | 99.79369 | 17371.53 | 1526.018 | 0 | 167.5382 | s(Year, k=10, bs="cr") + s(GDP, k=10, bs="cr") + s(Country, bs="re") |
| 8 | 3.855439 | 99.84861 | 16039.26 | 193.7514 | 8.46085812691349e-43 | 192.4324 | te(pbf.kcal, carb_prot.kcal, abf.kcal, Year, bs=c("tp", "cr"), d=c(3,1), k=c(k_nut, 7)) + s(Country, bs="re") |
| 9 | 3.855439 | 99.85481 | 15874.89 | 29.38286 | 4.1647851491474e-07 | 203.618 | te(pbf.kcal, carb_prot.kcal, abf.kcal, GDP, bs=c("tp", "cr"), d=c(3,1), k=c(k_nut, 7)) + s(Country, bs="re") |
| 10 | 3.855439 | 99.80421 | 17161.24 | 1315.727 | 1.96547598662957e-286 | 179.2737 | te(Year, GDP, k=10) + s(Country, bs="re") |
| 11 | 3.855439 | 99.85176 | 15957.29 | 111.7778 | 5.3425550952716e-25 | 198.3622 | te(pbf.kcal, carb_prot.kcal, abf.kcal, Year, bs=c("tp", "cr"), d=c(3,1), k=c(k_nut, 7)) + s(GDP, k=10, bs="cr") + s(Country, bs="re") |
| **12** | **3.855439** | **99.85586** | **15845.51** | **0** | **1** | **205.0184** | **te(pbf.kcal, carb_prot.kcal, abf.kcal, GDP, bs=c("tp", "cr"), d=c(3,1), k=c(k_nut, 7)) + s(Year, k=10, bs="cr") + s(Country, bs="re")** |
| 13 | 3.855439 | 99.80942 | 17060.2 | 1214.695 | 1.7074825617386e-264 | 188.9548 | te(Year, GDP, k=10) + s(pbf.kcal, carb_prot.kcal, abf.kcal, k=k_nut) + s(Country, bs="re") |

**Supplementary Table 4.** Estimated effects of macronutrient supply (plant- and animal-based fats and carbohydrate and protein) by time and GDP per capita on age-standardized ADHD incidence rate. Gaussian-GAMM, log-link function. (related to Figure 1G)

| **Parametric coefficients** | |  |  |  |
| --- | --- | --- | --- | --- |
|  | **Estimate** | **Std. Error** | **t value** | **Pr(>\|t\|)** |
| **(Intercept)** | 4.07357 | 0.04008 | 101.6 | <2e-16 |
| **Approximate significance of smooth terms** | | | |  |
|  | **edf** | **Ref.df** | **F** | **p-value** |
| **te(pbf.kcal,carb_prot.kcal,abf.kcal,GDP)** | 45.466 | 49.951 | 37.79 | <2e-16 |
| **s(Year)** | 1.583 | 1.964 | 16.52 | 8.41e-07 |
| **s(Country)** | 157.969 | 158.000 | 12081.77 | <2e-16 |
| R-sq.(adj) = 0.998 Deviance explained = 99.9% | | | | |
| GCV = 2.7456 Scale est. = 1.9454 n = 4465 | | | | |

**Supplementary Figure**

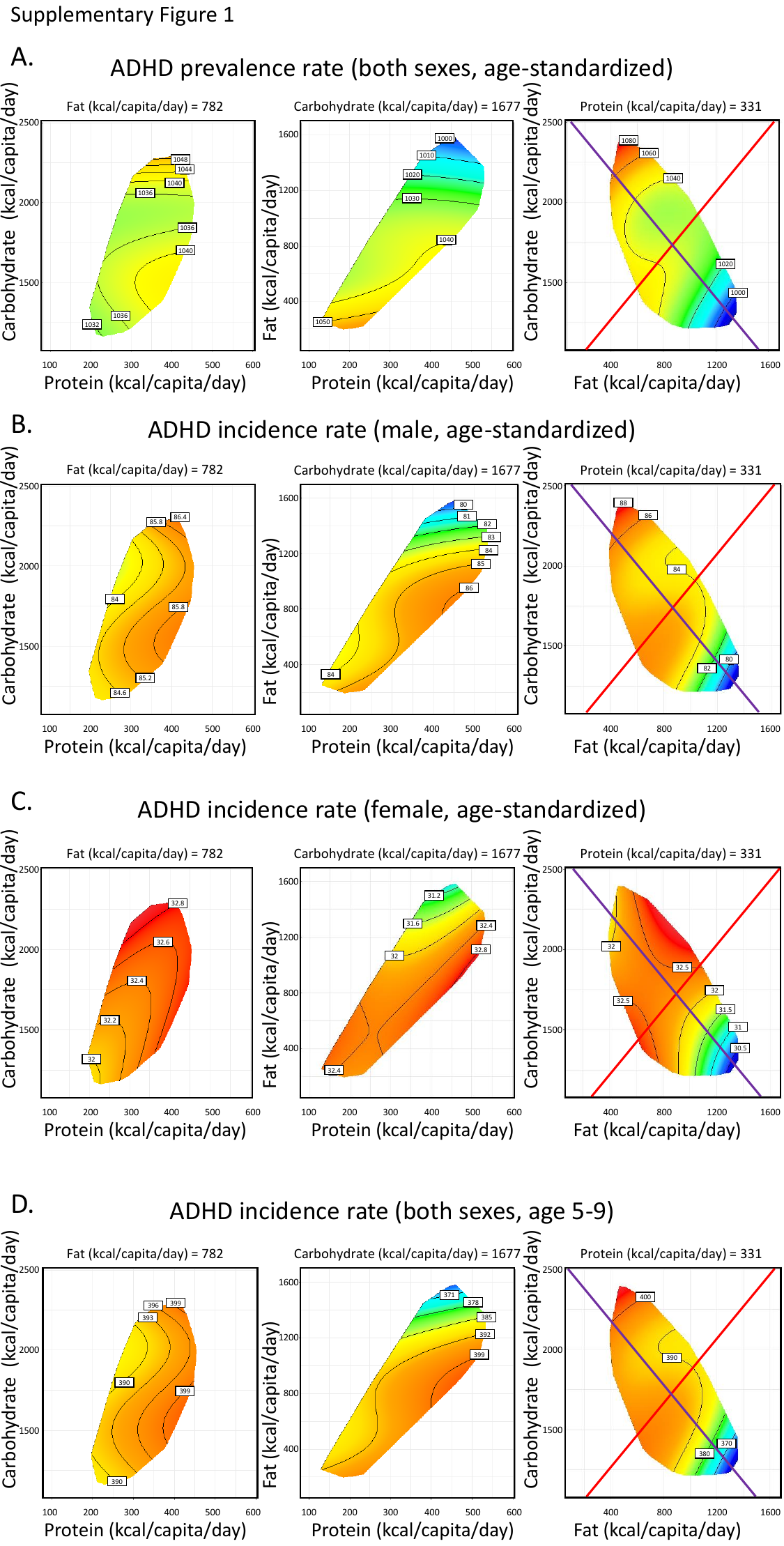

**Supplementary Figure 1.** Predicted effects of macronutrient supply on age-standardized ADHD prevalence rate of both sexes (**A**), age-standardized ADHD incidence rate of male (**B**), age-standardized ADHD incidence rate of female (**C**), and 5-9 years old ADHD incidence rate of both sexes (**D**).
